# Comorbidity Profiles of Posttraumatic Stress Disorder Across the Medical Phenome

**DOI:** 10.1101/2023.08.25.23294572

**Authors:** Emily M. Hicks, Maria Niarchou, Slavina Goleva, Dia Kabir, Julia Ciarcia, PTSD & Trauma EHR Working Group, Jordan W. Smoller, Lea K. Davis, Caroline M. Nievergelt, Karestan C. Koenen, Laura M. Huckins, Karmel W. Choi

## Abstract

**Background:** Prior epidemiological research has linked PTSD with specific physical health problems, but the comprehensive landscape of medical conditions associated with PTSD remains uncharacterized. Electronic health records (EHR) provide an opportunity to overcome prior clinical knowledge gaps and uncover associations with biological relevance that potentially vary by sex.

**Methods:** PTSD was defined among biobank participants (total N=123,365) in a major healthcare system using two ICD code-based definitions: broad (1+ PTSD or acute stress codes versus 0; N_Case_=14,899) and narrow (2+ PTSD codes versus 0; N_Case_=3,026). Using a phenome-wide association (PheWAS) design, we tested associations between each PTSD definition and all prevalent disease umbrella categories, i.e., phecodes. We also conducted sex-stratified PheWAS analyses including a sex-by-diagnosis interaction term in each logistic regression.

**Results:** A substantial number of phecodes were significantly associated with PTSD_Narrow_ (61%) and PTSD_Broad_ (83%). While top associations were shared between the two definitions, PTSD_Broad_ captured 334 additional phecodes not significantly associated with PTSD_Narrow_ and exhibited a wider range of significantly associated phecodes across various categories, including respiratory, genitourinary, and circulatory conditions. Sex differences were observed, in that PTSD_Broad_ was more strongly associated with osteoporosis, respiratory failure, hemorrhage, and pulmonary heart disease among male patients, and with urinary tract infection, acute pharyngitis, respiratory infections, and overweight among female patients.

**Conclusions:** This study provides valuable insights into a diverse range of comorbidities associated with PTSD, including both known and novel associations, while highlighting the influence of sex differences and the impact of defining PTSD using EHR.

## Introduction

Post-traumatic stress disorder (PTSD) occurs following highly stressful and/or traumatic events. Not only does PTSD impair occupational performance, social relationships, and overall quality of life, it frequently presents with comorbidities^1^, defined broadly here as co-occurring health conditions. Common PTSD comorbidities include psychiatric conditions such as depression and anxiety^2,3^, and physical health problems including cardiovascular disease^4^ and gastrointestinal conditions^5^. However, the comprehensive landscape of PTSD comorbidities, particularly in healthcare contexts, has remained incompletely characterized^6^. This limits clinical insights into complex needs of patients with PTSD (i.e., what other medical concerns to consider in formulating a treatment plan) as well as empirical insights into medical conditions that may serve as risk factors for PTSD and/or share underlying biological mechanisms^7^.

Electronic health records (EHR) provide a unique opportunity to uncover such associations with clinical and potential biological relevance. EHR offer large samples and longitudinal data including all diagnoses that a patient receives while active in that healthcare system, recorded using standardized diagnostic billing codes that can be examined in tandem across multiple systems. Such codes can be used in different ways to identify post-traumatic psychopathology^8^. While PTSD is designated by specific billing codes, providers may choose to assign a broader code to reflect diagnostic uncertainty, higher likelihood of insurance coverage, stigma reduction, or other factors, meaning that some PTSD cases may be missed when simply focusing on a narrow set of codes. In addition to identifying specific codes, we can also consider how frequently they are assigned to an individual. Accordingly, a broader definition might include a single diagnosis of any stress-related disorder versus a narrow definition requiring multiple diagnoses of PTSD specifically. This has implications for epidemiologic^9^ and genomic studies that leverage these definitions to identify PTSD cases and controls from EHR (for example, GWAS^10,11^), where a broader definition can inclusively capture more cases, enhancing discovery power, but may also introduce non-specific associations^12^. How we identify individuals with post-traumatic psychopathology using EHR, i.e., with more inclusive versus narrow definitions, may impact our ability to resolve relevant comorbidities, and the translatability of resulting genetic findings.

In addition to a need to resolve wide-ranging PTSD comorbidities, important sex differences may also exist in such comorbidities. The burden of PTSD varies by sex^3,13^, with rates of lifetime PTSD about twice as high in females compared to males^14^. Understanding why PTSD prevalence differs by sex is complex; rather than simply capturing intrinsic biological sex differences in risk, differences may reflect gendered environments including trauma exposure (e.g., interpersonal violence)^15–19^ or available supports. Male, female, and intersex patients may also have different experiences within healthcare systems, including where and how care is received, access to care, and their diagnostic journey for PTSD. As such, the medical conditions that co-occur with PTSD may also differ^20^.

In this multi-site study, we sought to investigate the relationships between different PTSD phenotypes and the medical phenome using data from 123,365 patients enrolled in biobanks across three major EHR-based healthcare systems. Our objectives were to comprehensively identify medical comorbidities associated with PTSD, and to examine whether these associations varied based on broad versus narrow definitions, and by sex. Finally, we explored how each PTSD definition relates to polygenic signals to understand implications for downstream genomic and clinical research.

## Methods

### Samples

Our study includes 123,365 biobank research participants with available genetic and longitudinal EHR data^21^ from three major health systems—Vanderbilt University Medical Center (VUMC, n=73,488), Mount Sinai (MSSM, n=31,704), and Mass General Brigham (MGB, n=29,885). Sample characteristics (**Table 1**) and phenotype category codes (phecodes) frequencies (**Supplementary Table 1**) and study information (**Supplementary Material 1**) for each site are provided.

**Table 1.**
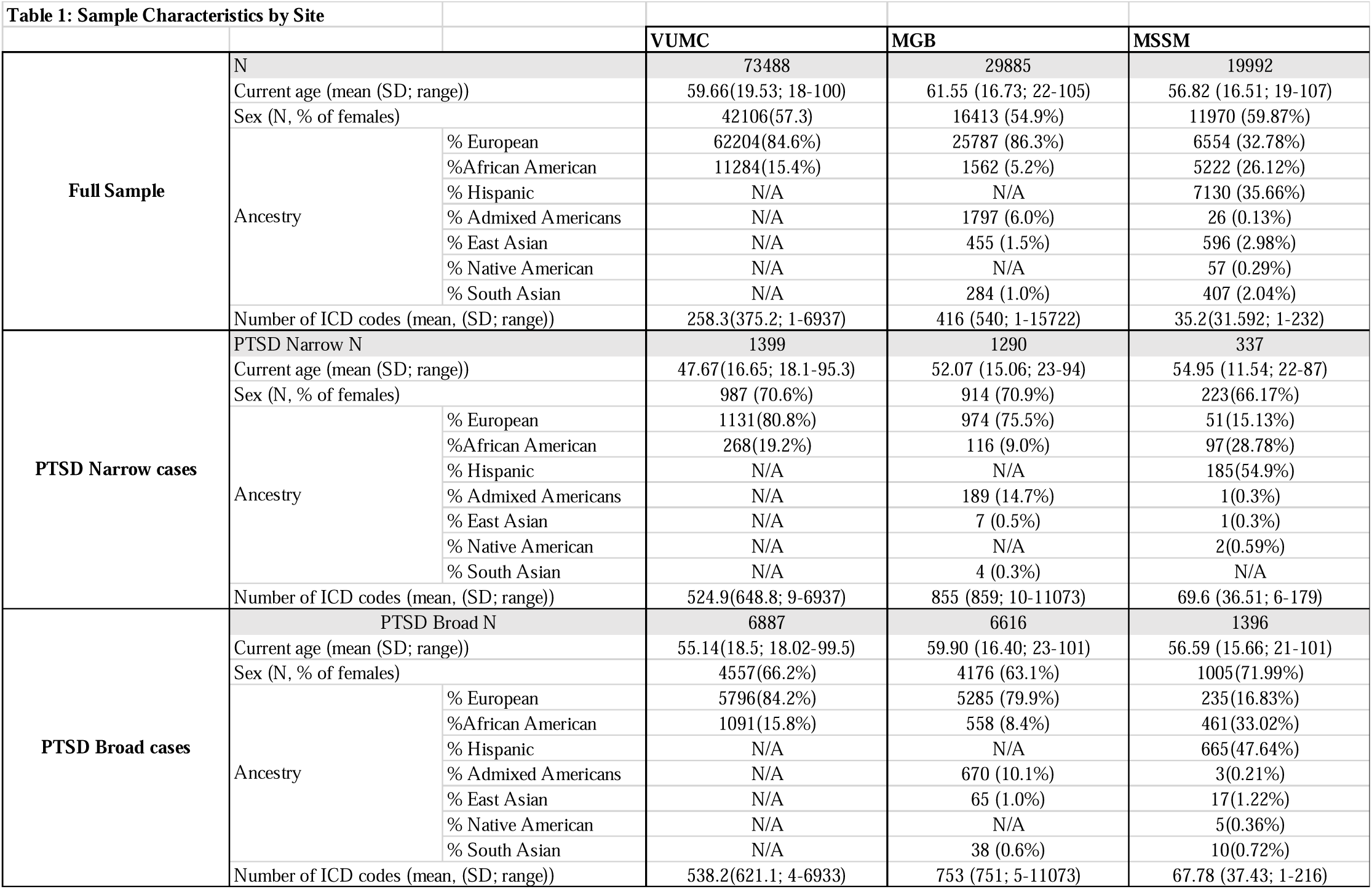
Sample characteristics by site.

## PTSD exposures

PTSD case-control phenotypes were defined using two ICD code-based definitions. Individuals who met broad PTSD criteria (PTSD_Broad_) had ≥ 1 PTSD (309.81, F43.1, F43.10, F43.11, F43.12) or stress-related disorder code (310.2, F07.81, 309, 309.1, 309.2, 309.21, 309.22, 309.23, 309.24, 309.28, 309.29, 309.3, 309.4, 309.82, 309.83, 309.89, 309.9, F43, F43.0, F43.2, F43.20, F43.21, F43.22, F43.23, F43.24, F43.25, F43.29, F43.8, F43.9), while controls had no such code; individuals who met narrow PTSD criteria (PTSD_Narrow_) had ≥ 2 PTSD codes only, while controls had no codes for PTSD, other stress-related disorders, or post-concussive injury.

## Main PheWAS analyses

We used a phenome-wide association (PheWAS) design to test logistic regression associations between PTSD_Narrow_ or PTSD_Broad_ with all phecodes as previously defined^22^. Cases for each outcome phecode had > 2 ICD codes belonging to that phecode category, while controls had 0 codes in that category (individuals with only one code were excluded). We excluded outcome phecodes derived from the same ICD codes used to define PTSD_Narrow_ (phecode 300.9 (“Posttraumatic Stress Disorder”)) and PTSD_Broad_ (phecodes 300.8 (“Acute Reaction to Stress”), 304 (“Adjustment Reaction”), and 300.9 (“Posttraumatic Stress Disorder”)) to eliminate circularity of exposure and outcome. We adjusted the models for age, genetically inferred ancestry (European [EUR], African [AA], Americas [AMR], East Asian [EAS], where available at each site), sex, and top 5 PCs to adjust for residual differences. As a sensitivity analysis, each site also tested PheWAS models in which genetic PCs were not included as a covariate. We applied a Bonferroni threshold to establish significance, correcting for 1324 tests (pBONF<3.7×10^−5^; PTSD_Narrow_) and 1367 tests (pBONF<3.8×10^−5^; PTSD_Broad_). We assessed enrichment of phecode categories of associations reaching Bonferroni significance, and the top 25% of associations, using a binomial test.

For all phecodes appearing in ≥ 2 sites in the analysis (1649 phecodes were available for ≥2 sites; 765 were available for all three sites), we meta-analyzed across sites using fixed-effects in the *PheWAS* R package^23^. To assess heterogeneity, we calculated I^2^ statistics in METAL^24^ to identify associations that differed in effect sizes between health systems. Phecodes with I^2^<0.1 were considered significantly heterogeneous. While we observed significant heterogeneity among effect size estimates across sites for both PTSD_Narrow_ and PTSD_Broad_ PheWAS, directions of effect and top associations across sites remained largely uniform (**Supplementary Table 2**).

## Sex-stratified and sex-interaction PheWAS Analyses

We conducted sex-stratified PheWAS analyses (n=13,228 female, n=11,614 male), adjusting for age, ancestry, and top 5 PCs. Patient sex was categorized as follows: MGB: sex assigned at birth, VUMC: sex assigned at birth, Mount Sinai: karyotypic sex; a small number of intersex individuals and individuals who declined to state their sex were excluded from this analysis. For all Bonferroni-significant phecodes from the full or sex-stratified analyses, we conducted an analysis including a sex-by-diagnosis interaction term in each logistic regression. Interaction effect sizes were meta-analyzed using an inverse-variance weighted model using a “STDERR” scheme with METAL software^24^.

## Polygenic score analyses

We assessed genetic correlates of PTSD definitions in individuals of European (EUR) and African ancestries (AA) based on PTSD polygenic scores (PGS). We calculated PGS inferring SNPs weights from European PGC-PTSD GWAS summary statistics (discovery n=174,659)^25^ as implemented in PRS-CSx^26^ and summing allele counts weights. For each available ancestry group within each health system, we normalized the PGS and tested their association with PTSD_Narrow_ or PTSD_Broad_ using logistic regression, including sex, age, and within-ancestry genotype-derived principal components (PCs). We estimated phenotypic variance explained by PTSD scores using Nagelkerke’s R^2^,^27^. We meta-analyzed PGS associations with fixed effects using the metafor package^28^.

## Results

### Sample

To examine phenome-wide comorbidities of PTSD, we conducted analyses included 123,365 patients (3,026 and 14,899 meeting the PTSD_Narrow_ versus PTSD_Broad_ definitions, respectively) across three major healthcare systems: VUMC, MGB and MSSM (**Table 1**). Prevalence of PTSD_Narrow_ and PTSD_Broad_ at MGB (4.32% and 22.14%, respectively) were higher than the other two sites (VUMC: 1.9% and 9.37%, MSSM: 1.69% and 6.98%).

## Narrowly and broadly defined PTSD definitions have wide-ranging correlates across the medical phenome

807 of 1324 (61%) phecodes were significantly associated with PTSD_Narrow_ and 1136 or 1367 (83%) with PTSD_Broad_ (p_Bonf_<3.8×10^−5^, 3.7×10^−5^, respectively; full summary statistics: **Supplementary Table 2**). Top PTSD_Narrow_ associations included psychiatric and pain-related phecodes, whereas top PTSD_Broad_ associations included psychiatric and pain-related phecodes as well as respiratory, genitourinary, and circulatory phecodes (**Figure 1**). Phecode associations with PTSD were primarily positive: we observed an increased risk of each of these phecodes among individuals with PTSD. Only a very small minority of phecodes was negatively associated with PTSD, including regional enteritis (Crohn’s) (p=0.019, OR=0.77; p=0.0061, OR=0.86 for PTSD_Narrow_ and PTSD_Broad_, respectively) and prostate cancer (p=0.014, OR=0.61; p=0.022, OR=0.87) (**Supplementary Table 2**). Notably, these negative directions of effect were consistent across all sites.

**Figure 1.**
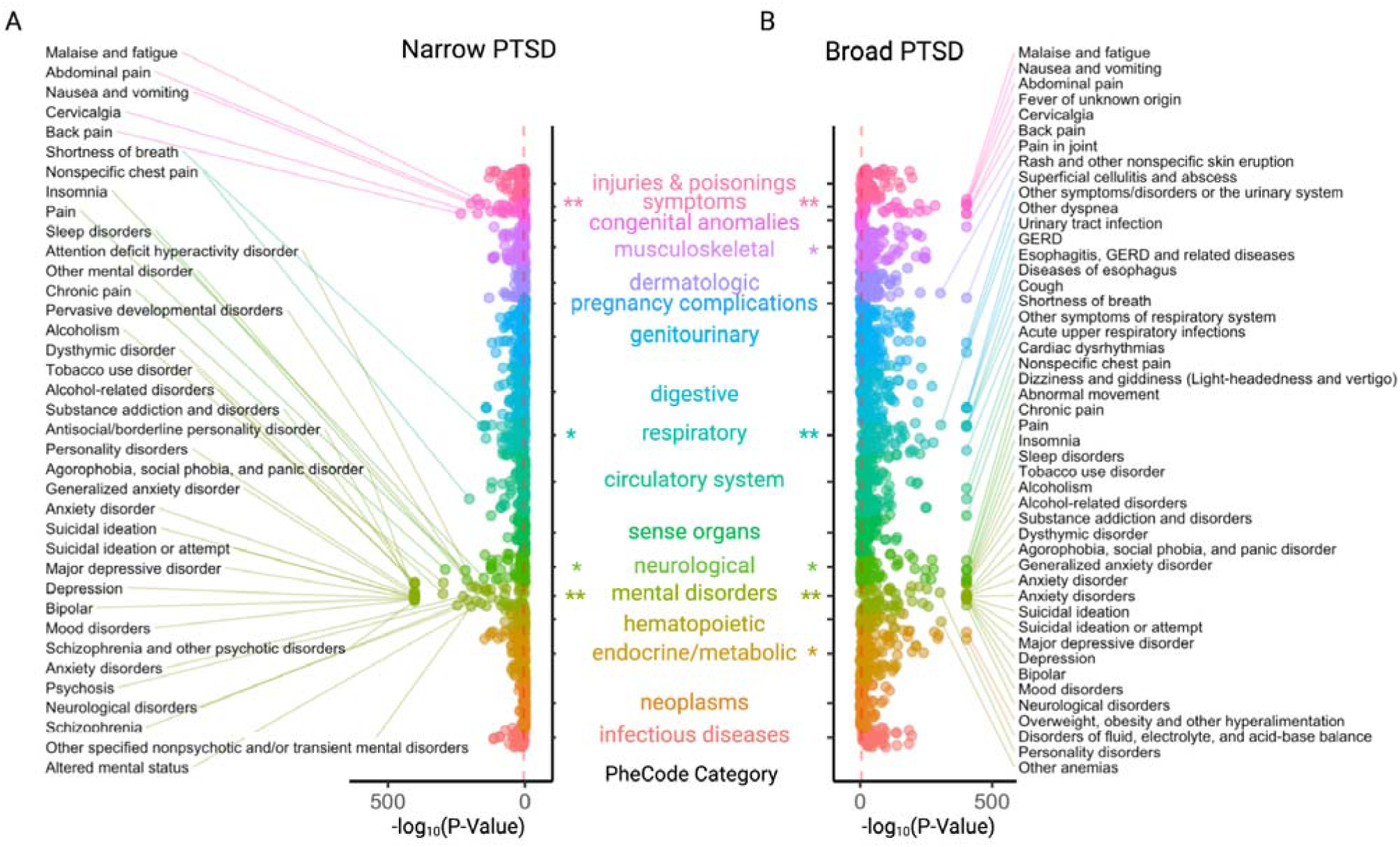
A) PheWAS meta-analysis results for the narrow PTSD definition. B) PheWAS meta-analysis results for the broad PTSD definition. Phecodes are organized across the y-axis by their category, with the strength of association shown in the x-axis using –log10p. Red dashed line indicates the Bonferroni significance threshold. Bonferroni (**) and nominally significant (*) phecode category enrichments for the top 25% of phecode associations for PTSD_Narrow_ (mental disorders: p<0.0001, symptoms: p=0.0001, neurological: p=0.0019, respiratory=0.0081) and PTSD_Broad_ (mental disorders: p<0.0001, symptoms: p<0.0001, respiratory: p=0.0007, neurological: p=0.0269, musculoskeletal: p=0.0324, endocrine/metabolic: p=0.0312)

## Narrowly-defined PTSD shows strong associations with neuropsychiatric traits

PTSD_Narrow_ was associated with over 85% of phecodes in the mental disorders category (**Supplementary Table 3**). Among the top 25% of phecodes associated with PTSD_Narrow_, ‘mental disorders’ and ‘symptoms’ categories were significantly enriched after Bonferroni correction (**Supplementary Table 3**). The strongest associations with PTSD_Narrow_ fell in the mental disorders category, including personality disorders (*Antisocial/borderline personality disorder, Personality disorders*); mood disorders (*Mood disorders, Depression*); suicide-related phecodes (*Suicidal ideation, Suicidal ideation or attempt*); substance use disorders (*Substance addiction and disorders, Alcohol-related disorders*); and anxiety disorders (*Agorophobia, social phobia, and panic disorder, Anxiety disorder, Generalized anxiety disorder*). Beyond the mental disorders category, PTSD_Narrow_ was associated with over 85% of phecodes in the infectious diseases category (**Supplementary Table 3**). Phecodes with the largest odds ratios in association with PTSD_Narrow_ were *Anxiety Disorders*, *Antisocial/borderline personality disorder*, *personality disorder*, *mood disorder*, *suicidal ideation*, *suicide or self-inflicted injury*, *poisoning by psychotropic agents,* and *depression*.

Additionally, PTSD_Narrow_ showed strong associations with phecodes in the neurological and symptoms categories pertaining to pain, including *Chronic pain, Back pain, Pain*, and *Nonspecific chest pain*. Several genitourinary phecodes also emerged in association with PTSD_Narrow_, for example, *Other symptoms/disorders of the urinary system*, *Urinary tract infection*, and *Dysuria*.

## Broadly-defined PTSD shows strong associations with neuropsychiatric traits and respiratory, digestive, genitourinary, and cardiovascular conditions

PTSD_Broad_ was significantly associated with all phecodes in the mental disorders and infectious disease categories and over 90% of phecodes in the respiratory, injuries & poisonings, symptoms and digestive categories (**Supplementary Table 3**). Among the top 25% of significant phecodes, mental disorders, symptoms, and respiratory PheCode categories were significantly enriched (**Supplementary Table 3**). The top phecode associations with PTSD_Broad_ included mood disorders (*depression, suicidal ideation, anxiety, bipolar disorder*), substance use disorders (*Substance addiction and disorders, alcohol related disorders alcoholism, tobacco use disorder*), and other psychiatric disorders (*schizophrenia, ADHD*). The phecodes with the largest odds ratios in association with PTSD_Broad_ were *antisocial/borderline personality* (OR=18.8 (18.61-19.00)), *personality disorder* (OR=13.21 (13.07-13.34)), *suicidal ideation* (OR=9.4 (9.30-9.51)), and *anxiety disorders* (OR=9.37 (9.33-9.41)).

In addition to mental disorders, top associations with PTSD_Broad_ were related to a wide range of medical phecode categories, including sleep (*insomnia, sleep disorders, malaise and fatigue*), pain (*back pain, chronic pain, abdominal pain, pain, pain in joint*), respiratory system (*other symptoms of respiratory system, cough, acute upper respiratory infections of multiple or unspecified sites, shortness of breath, pneumonia*), digestive system (*nausea and vomiting, GERD, diseases of esophagus, esophagitis*), genitourinary system (*urinary tract infection, other symptoms disorders of the urinary system, dysuria*), cardiovascular system (*nonspecific chest pain, cardiac dysrhythmias, hypertension, hypotension, arrhythmia, palpitations*), and asthma conditions.

## Clinical phenome associations were largely shared but showed some differences across PTSD definitions

Significant phecode associations were largely shared between PTSD_Narrow_ and PTSD_Broad_ definitions, especially the top-ranking associations (**Figure 2**). However, many more phecodes were significantly associated with PTSD_Broad_ (1136/1367 tested) compared to PTSD_Narow_ (807/1324), and 334 Bonferroni-significant phecodes were associated with PTSD_Broad_ but not PTSD_Narrow_. The strongest such associations included phecodes related to skin (neoplasms of skin, dyschromia, actinic keratosis, other non-epithelial cancer of skin, dermatitis), lung transplant and chronic pulmonary heart disease; however, these phecodes were not among the strongest hits for PTSD_Broad_. Only four phecodes were significantly associated following Bonferroni correction with PTSD_Narrow_ but not PTSD_Broad_. These included relatively uncommon genitourinary phecodes such as urethral hypermobility/ISD, pelvic peritoneal adhesions, as well as amblyopia, and other disorders of middle ear and mastoid.

**Figure 2.**
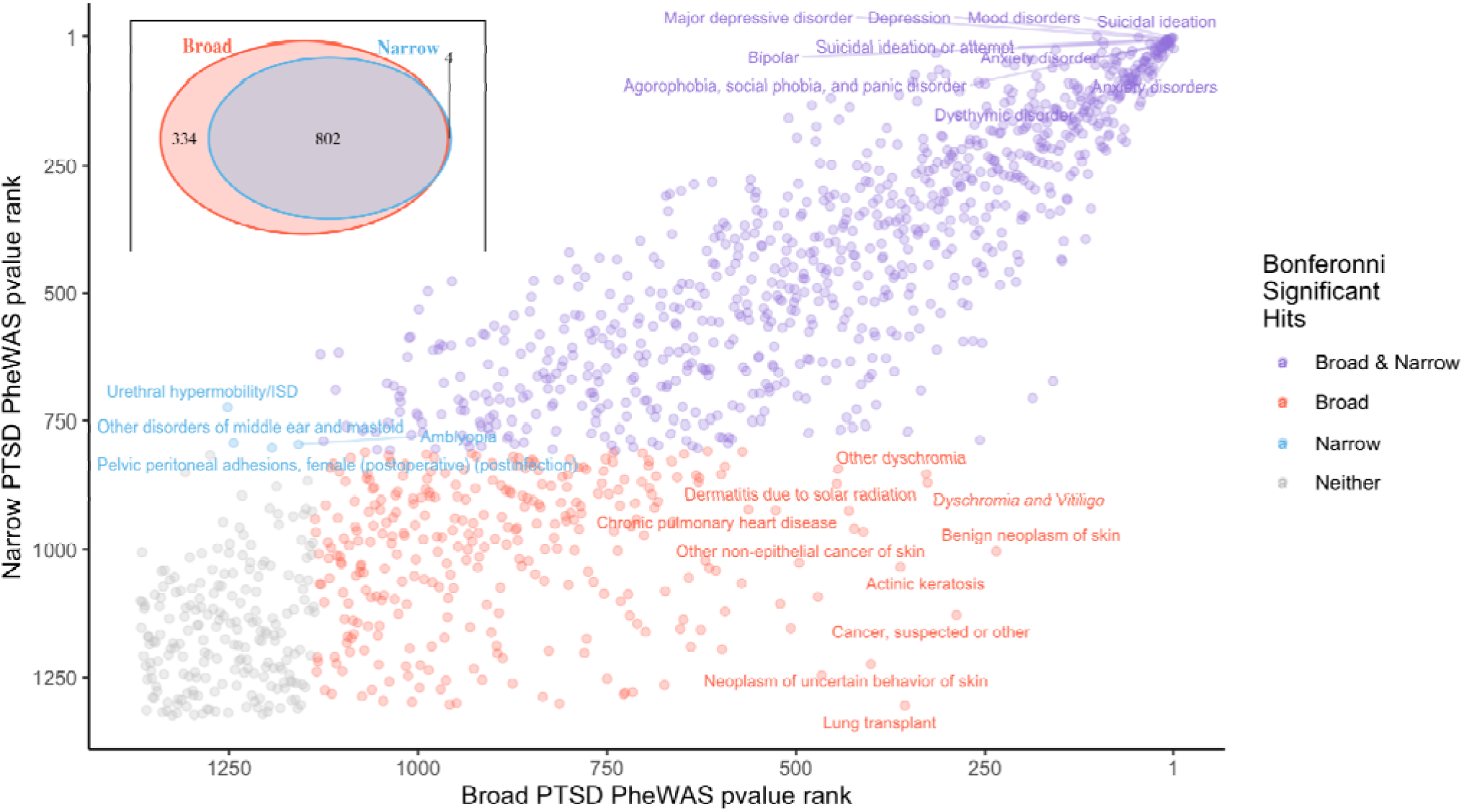
Top PheWAS hits are shared between narrowly and broadly defined PTSD. Phecodes ranked by p value are plotted for PTSD_Broad_ (x-axis) and PTSD_Narrow_(y-axis). Phecodes with Bonferroni significant association with PTSD_Narrow_ (blue), PTSD_Broad_ (red), both (purple) or neither (gray) are indicated. Number of shared and distinct phecode associations for PTSD_Narrow_ and PTSD_Broad_ are shown in the inset.

## Sex moderates associations between broadly-defined PTSD and several other phenotypes

We conducted sex-stratified PTSD_Narrow_ and PTSD_Broad_ PheWAS, and meta-analyzed across sites (**Supplementary figures 1-2; Supplementary Table 4**), and performed a PTSD-by-sex interaction test restricted to only those phecodes with a Bonferroni-significant main effect within either sex (**Figure 3**). There were 16 phecodes that were significantly different between males and females coded with PTSD_Broad_ after meta-analysis. Phenotypes that were more common among male patients with PTSD_Broad_ included osteoporosis, respiratory failure, subarachnoid and intracranial hemorrhage, and pulmonary heart disease. Phenotypes that were more common among female patients with PTSD_Broad_ included urinary tract infection and other urinary symptoms, acute pharyngitis, acute respiratory infections, and overweight, though directions of effect were not consistent between sites for some phecodes. For PTSD_Narrow_, other symptoms/disorder of the urinary system was the only phecode with a significant sex interaction, driven by higher prevalence in males at MGB despite opposing associations observed at the other two sites.

**Figure 3.**
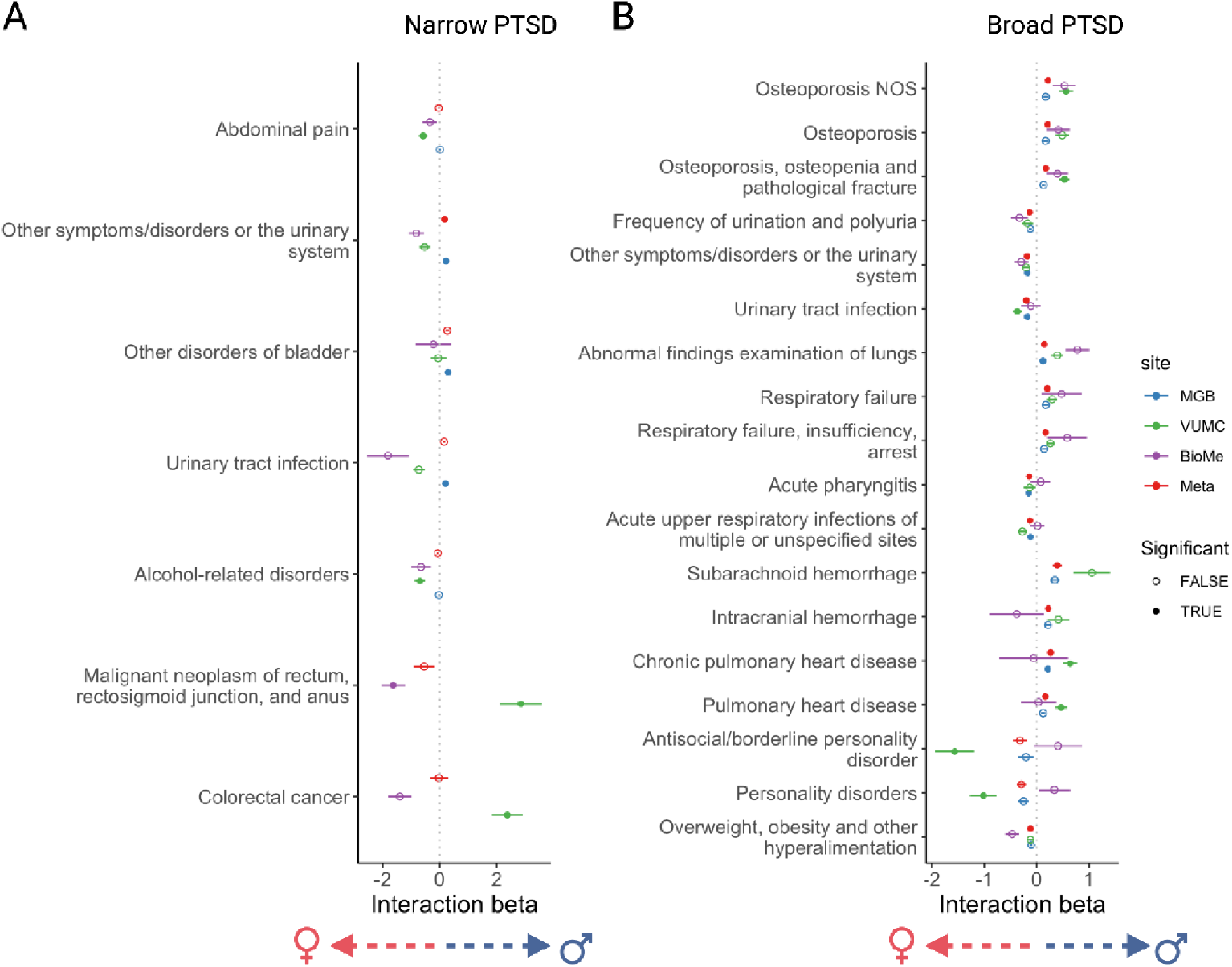
Phecodes with significant sex interaction with PTSD_Narrow_ (A) and PTSD_Broad_ (B) in at least one site or in meta-analysis across sites. Interaction term (PTSD * Sex) coefficients and standard errors are plotted for each site independently (MGB=blue, VUMC=green, MSSM=purple) and for a meta-analyzed interaction term coefficient (red). Bonferonni significant interaction terms are indicated with a filled circle. Positive interaction term indicates a higher prevalence in males and a negative interaction term indicates a higher prevalence in females.

## Explanatory power of polygenic scores varies according to PTSD definition

We tested PGS associations with both PTSD_Narrow_ and PTSD_Broad_ (**Table 2)**. PTSD PGS significantly associated with both PTSD_Narrow_ (meta-analytic B=0.17, SE=0.02, p=5.6E-14, R^2^ range= 0.08-0.29%; total cases: 11,075) and PTSD_Broad_ (meta-analytic B=0.07, SE=0.01, p=2.7E-11, R^2^ range=0.08-0.43%; total cases: 2,116) in EUR subsamples, although the proportion of variance explained (R^2^) was low. PGS were not significantly associated among AA individuals for either PTSD_Narrow_ (B=0.06, SE=0.09, p= 0.30, R^2^ range=0.0002-0.04%) or PTSD_Broad_(B=0.01, SE=0.09, p=0.53, R^2^ range=0.006-0.32%), likely due to poor representation of African ancestry alleles in existing PTSD GWAS.

**Table 2.** PTSD polygenic score (PGS) Meta-analysis association statistics.

| Table 2: PGS Meta-analysis association statistics |  |  | PTSD ~ PRS + Age + Sex + PCs1:5 |  |  | where PRS is scaled |  |  |  |
| --- | --- | --- | --- | --- | --- | --- | --- | --- | --- |
| Ancestry Group | PTSD Definition | Site | Beta | se | p | N case | N control | N total | Nagelkerke |
| EUR | Narrow | MGB | 0.1951 | 0.0344 | 0.00000008 | 936 | 19591 | 20527 | 0.425% |
|  |  | VUMC | 0.1500 | 0.0300 | 0.00000179 | 1131 | 56168 | 57299 | 0.230% |
|  |  | BioMe | 0.1117 | 0.1719 | 0.51576118 | 49 | 6307 | 6356 | 0.078% |
|  |  | META-ANALYSIS | 0.1685 | 0.0224 | 5.6E-14 |  |  |  |  |
|  | Broad | MGB | 0.0653 | 0.0161 | 0.0000476 | 5059 | 19765 | 25698 | 0.081% |
|  |  | VUMC | 0.0700 | 0.0140 | 0.0000018 | 5796 | 56408 | 62204 | 0.083% |
|  |  | BioMe | 0.1800 | 0.0824 | 0.0290112 | 220 | 6137 | 6357 | 0.291% |
|  |  | META-ANALYSIS | 0.0698 | 0.0105 | 2.7E-11 |  |  |  |  |
| AA | Narrow | MGB | - | - | - | - | - | - | - |
|  |  | VUMC | 0.0250 | 0.0700 | 0.7226 | 268 | 10158 | 10426 | 0.006% |
|  |  | BioMe | -0.1723 | 0.1041 | 0.0980 | 96 | 5062 | 5158 | 0.315% |
|  |  | META-ANALYSIS | -0.0364 | 0.0581 | 0.5305 |  |  |  |  |
|  | Broad | MGB | - | - | - | - | - | - | - |
|  |  | VUMC | 0.0500 | 0.0370 | 0.1600 | 1091 | 10193 | 11284 | 0.038% |
|  |  | BioMe | -0.0037 | 0.0496 | 0.9404 | 458 | 4703 | 5161 | 0.000% |
|  |  | META-ANALYSIS | 0.0308 | 0.0297 | 0.2990 |  |  |  |  |

## Discussion

This study examined phenome-wide relationships between EHR-based PTSD and over 1300 medical phecodes among over 123,000 participants across 3 health systems, including 14,899 and 3,026 individuals meeting broad and narrow definitions for PTSD, respectively. We identified known and novel PTSD comorbidities that replicated across multiple health systems and contextualized the relative strength of PTSD-related associations across the medical phenome. Additionally, we identified several phenotypes for which males and females exhibit differing relationships with PTSD. These results characterize a wide range of PTSD-related diagnostic comorbidities in a health system setting and provide a useful catalog of phenotype-phenotype relationships for interpreting future biobank-based genetic association studies of PTSD.

## Contextualizing known and novel PTSD comorbidities

Our results build upon known psychiatric^29–31^ and non-psychiatric PTSD comorbidities. Psychiatric disorders were consistently identified as the strongest comorbidities for both broad and narrow PTSD definitions; among non-psychiatric disorders, we observed a strong association of both PTSD definitions with pain phenotypes, which encompass physical sensory components, as well as cognitive and emotional components^32–36^. Given inflammatory/immune links to PTSD^37^, we were surprised to find that autoimmune disorders such as rheumatoid arthritis and multiple sclerosis were not significantly associated with PTSD. This suggests these comorbidities may be less common on a health system scale, or that these comorbidities present with specific PTSD subtypes (for example, those with a specific immune component or etiopathology) that are not well represented or captured in our biobanks. Unexpectedly, we did not find significant associations between PTSD and decreased libido^38^. However, this phecode is relatively scarce in our biobanks (N=162-214), decreasing our ability to detect associations. We identified novel associations of genitourinary phenotypes with PTSD, with a particularly strong association with PTSD_Broad,_ with significance levels comparable to respiratory and cardiovascular comorbidities. It is possible that these associations stem from shared risk factors, i.e., that sexual assault or related traumatic experiences lead to both genitourinary and psychiatric outcomes^39^, or are artefactual associations resulting from cross-disorder correlations, where conditions that appear significant are not directly associated with the exposure but rather linked to other diseases. It is also possible these associations reflect “healthcare pleiotropy” – i.e., the association stems from an artifact of the clinical settings where traumatic exposures are more commonly/routinely discussed (i.e., OB/GYN clinics).

## Sex differences in PTSD comorbidities

Epidemiological evidence indicates that females exposed to traumatic events are at higher risk for developing PTSD compared to trauma-exposed males. We identified several phenotypes for which associations with PTSD_Broad_ differed between males and females. For females, obesity had a stronger association with PTSD than males. This relationship has been previously reported for some studies of PTSD^40,41^ and other psychiatric phenotypes^42^. We also found stronger association of urinary symptoms with PTSD in females. This may be due to genito-urinary symptoms that occur at higher rates following exposure to abuse^38^; for example, interstitial cystitis/bladder pain syndrome^43^, vulvodynia^44^, and chronic pelvic pain^45,46^, coupled with a greater prevalence of sexual trauma among females^17,47^ or perhaps due to the clinical setting where trauma and genitourinary symptoms are assessed. Respiratory failure, cranial hemorrhage, and pulmonary heart disease were more prevalent among male PTSD_Broad_ cases. Together these results may reflect differential patterns of diagnosis of these disorders or PTSD, differences in the care received between sexes (for example, in medical fields with sex specificity like gynecology) or sex differences in PTSD pathology and the downstream system-wide consequences of having PTSD.

## Scope of PTSD phenotyping and implications for genetic studies

This study explored the role of phenotypic specificity in resolving comorbid phenotypes. A narrow definition requiring multiple PTSD codes likely captures individuals with more severe and specific symptomatology. In contrast, the broad definition, consisting of a single code inclusive of stress-related reactions, may include individuals with a broader range of symptom severity. Psychiatric genetic studies increasingly tend to include broader definitions which facilitate greater sample sizes, but at the potential cost of specificity^12^. Both PTSD definitions showed strikingly high levels of comorbidities. However, we found top PTSD_Narrow_ associations were concentrated in mental disorders and pain-related symptoms, whereas PTSD_Broad_ associations were spread across more phecode categories such as respiratory, genitourinary and circulatory conditions. Generally, the top phecodes were consistent across both definitions, with the broader definition associated with 300 additional phenotypes.

These association patterns have significant implications for our understanding of broad vs. narrow definitions of psychiatric disorders and may shape future analytical and inclusion/exclusion criteria, as well as how we should interpret associations specific to PTSD_Broad_. The most significant of these were skin conditions (dyschromia, actinic keratosis etc.), which may reflect racial differences in PTSD prevalence, trauma exposure, or diagnostic rates^15^. To assess relevance for future studies, we consider each definition. If PTSD_Broad_ captures both full PTSD and sub-threshold PTSD cases (or, if an accurate diagnosis is obtained using only one ICD code, PTSD_Broad_ may capture primarily full PTSD cases), then we might conceptualize these associations as true PTSD associations, discoverable only thanks to this increase in power. However, if the broad category involves decreased diagnostic accuracy, PTSD_Broad_ may simply add ‘noise’ to the analysis, rather than identifying sub-threshold or otherwise ‘missed’ cases, and these additional associations may not contribute meaningfully to our understanding of PTSD and its comorbidities. At the genetic level, we observed that a PTSD PGS (trained on a GWAS based on mainly studies of clinically ascertained PTSD^25^) significantly associated with both PTSD_Broad_ and PTSD_Narrow_ status, though more strongly PTSD_Narrow_ than PTSD_Broad_. In the future, as sample sizes for PTSD GWAS increase, focusing on more strictly defined cases may become more feasible and may lead to increased predictive power of PGS.

Several factors suggest that the former, rather than latter, conceptualization is correct. First, addition of truly random cases (i.e., introduction of ‘noise’) should reduce PheWAS discovery power^48^ rather than identify additional significant associations. Second, both PTSD_Broad_ and PTSD_Narrow_ have significant psychiatric associations; it does not appear that one group is obviously more ‘accurately’ ascertained that the other. Third, we note a primary difference between the two groups reflects frequency of diagnosis; a single ICD code for PTSD does not necessarily imply an incorrect diagnosis, sub-clinical presentation, or less severe symptoms (this could be examined through in-depth chart reviews in the future); rather, these patients may leave the medical system or be referred elsewhere for treatment. In particular, we note that MSSM is associated with a VA, where veterans experiencing combat-related PTSD may pursue PTSD care instead of the initial diagnostic site.

## Strengths and limitations

Our study has several limitations to note when assessing and interpreting our results. First, while studying the full clinical phenome using EHR is a strength of our study, our biobank-based patient populations may not be fully representative of the general population. To some extent, we mitigate this concern by comparing effect sizes and meta-analyzing^48^ across three healthcare settings that represent distinct geographical areas (Boston, New York, Nashville), with differing demographics and healthcare needs (e.g., the proportion of veterans seeking care at these clinics differs significantly), and differing proportions of individuals with PTSD. In particular, one site (MGB) has a much higher rate of PTSD diagnosis (PTSD_Narrow_: 4.3% vs 1.9%, 1.7% at VUMC, MSSM, respectively). Consequently, we observe some heterogeneity in effect size of associations; however, effect directions are largely consistent across all three sites, which is highly unlikely to be due to chance (655 concordant across three sites vs. 169 discordant: binomial p=4.84×10^−236^; for codes present in two sites only: 340 concordant vs. 126 discordant, binomial p=9.06×10^−24^).

Given the majority of individuals are seeking care in a hospital setting for some symptom or condition, EHR-based analyses likely underestimate the proportion of healthy individuals and will not identify sub-threshold PTSD or those with well-managed PTSD receiving care in a different system. Our analysis also does not adjust for utilization rates. Relatedly, individuals in our study may have uneven access to healthcare or to certain specialists within a healthcare system: psychiatric or mental health services in particular may be unevenly accessed due to differences in insurance coverage or stigma or immigration status. Even when healthcare is accessed, patients may receive differing levels of care and stereotyping bias or institutionalized racism in healthcare may lead to under-or over-diagnosis^48–50^. These issues may be particularly exacerbated in psychiatry, where certain presentations of PTSD may be more readily diagnosed, potentially biasing our comorbidity results towards that subtype of PTSD.

Another limitation is the lack of temporal resolution. Understanding whether these associated phecodes occur prior to, or following, PTSD onset would ideally allow us to delineate associations into antecedents versus potential consequences of PTSD diagnosis. However, this is difficult in practice: patients may report longstanding existing diagnoses to a clinician upon enrollment at a new clinic, or receive correct diagnoses after long diagnostic journeys. In the latter case, a patient may experience initial symptoms several years before an accurate diagnosis of PTSD, such that other diagnoses received may either be comorbid with PTSD or represent inaccurate diagnoses given to explain some of the PTSD symptoms (for example, MDD, or adjustment reaction disorder). Difficulty in pinpointing disease onset precludes analysis of temporal relationships. Informed by this comprehensive map of associations, studies should be repeated in longitudinal cohorts with detailed and repeated psychiatric questionnaires that provide nuanced temporal resolution and can help shed light on the temporal nature of these phenotype-phenotype relationships in PTSD. Further exploration of the genetic factors contributing to PTSD and its comorbidities is also warranted.

## Conclusions and future directions

This multi-site study investigated relationships between PTSD phenotypes and medical comorbidities using data from over 123,000 patients across three major EHR-based healthcare systems. Our findings provide insights into the diagnostic complexity of PTSD and its associations with the clinical phenome. By considering both broad and narrow PTSD definitions, we comprehensively identified known and novel diagnostic comorbidities, particularly in the domains of mental disorders, pain, respiratory, genitourinary, and cardiovascular conditions. These associations were observed across multiple health systems, highlighting their generalizability. Importantly, we observed sex differences in the associations between PTSD and certain medical phenotypes, underscoring the need to consider sex-specific factors. Furthermore, our genetic analyses revealed significant associations between polygenic scores derived from GWAS and both broad and narrow definitions of PTSD. Future research should include longitudinal analyses to examine temporal relationships between PTSD and medical comorbidities, providing insights into the directionality of these associations. Investigating sex-specific factors, such as hormonal influences, trauma response differences, or care-seeking patterns, will enhance our understanding of sex disparities in PTSD and associated medical conditions, while mechanistic studies could explore underlying biological pathways and shared genetic factors contributing to the observed associations. Overall, this study deepens our understanding of the diagnostic complexity of PTSD within EHRs and provides insights for research towards integrated treatment approaches that address both PTSD diagnosis and its comorbidities.

## Acknowledgements

This collaborative work was supported in part by the PsycheMERGE grant MH118233 (to JWS and LKD) and administrative supplement to PGC PTSD R01 (R01MH106595). KWC was supported in part by funding from the National Institute of Mental Health (K08MH127413) and a NARSAD Brain and Behavior Foundation Young Investigator Award. EMH, JC and LMH acknowledge funding from NIMH (R01MH124839, R01MH118278) and NIEHS (R01ES033630). LKD was supported by NIMH (R01MH118223); NHGRI and the Office of Research on Women’s Health (RM1HG009034).

MSSM: This work was supported in part through the computational resources and staff expertise provided by Scientific Computing at the Icahn School of Medicine at Mount Sinai under award number S10OD018522 from the Office of Research Infrastructure of the National Institutes of Health. This work was supported through the resources and staff expertise provided by the Charles Bronfman Institute for Personalized Medicine and the Bio*Me* Biobank Program at the Icahn School of Medicine at Mount Sinai.

BioVU: The dataset(s) used for the analyses described were obtained from Vanderbilt University Medical Center’s BioVU which is supported by numerous sources: institutional funding, private agencies, and federal grants. These include the NIH funded Shared Instrumentation Grant S10RR025141; and CTSA grants UL1TR002243, UL1TR000445, and UL1RR024975. Genomic data are also supported by investigator-led projects that include U01HG004798, R01NS032830, RC2GM092618, P50GM115305, U01HG006378, U19HL065962, R01HD074711; and additional funding sources listed at https://victr.vumc.org/biovu-funding/. This publication was supported by Grant No. R01 HG011405, partially funded by the Office of Research on Women’s Health, Office of the Director, NIH and the National Human Genome Research Institute. Its contents are solely the responsibility of the authors and do not necessarily represent the official views of the Office of Research on Women’s Health or the National Human Genome Research Institute.

Disclosures: JWS is a member of the Scientific Advisory Board of Sensorium Therapeutics (with equity), and has received grant support from Biogen, Inc. He is PI of a collaborative study of the genetics of depression and bipolar disorder sponsored by 23andMe for which 23andMe provides analysis time as in-kind support but no payments.

## Supplementary Tables

1. Rank of top phecodes by site
2. Full summary statistics for PTSD PheWAS
3. PTSD PheWAS hits by PheCode category and Cross-category enrichment of PheCode categories for PTSD PheWAS hits
4. Full summary statistics for sex-stratified PTSD PheWAS

## Supplementary Figures

1. Male and female stratified PheWAS plot for PTSD Narrow
2. Male and female stratified PheWAS plot for PTSD Broad.

## Data Availability

All data produced in the present work are contained in the manuscript and supplemental materials.

