## Supplemental Materials for "Comorbidity Profiles of Posttraumatic Stress Disorder Across the Medical Phenome"

**VUMC’s BioVu.** Diagnostic codes from the International Classification of Disease versions 9 or 10 (ICD-9/10), were obtained from VUMC’s EHR database of >3.2 million individuals. These data are linked to VUMC’s biobank, BioVU, with DNA collected from >250,000 individuals. The VUMC Institutional Review Board (IRB) approved this study (protocol #190418), and all participants provided written informed consent. At the time of analysis, genotype data were available for 73,488 unrelated individuals of European and African ancestry. Details on the quality control procedures are described elsewhere^22^.

**MSSM’s Bio*Me*.** ICD-9/10 codes from MSSM’s EHR database were obtained for 31,704 individuals enrolled in the Bio*Me* biobank. Bio*Me* enrollment is unrestricted (non-selective in terms of gender, race, ethnicity, age, medical condition, or disease status) and all participants provide genetic data derived from blood samples at the time of enrollment. BioMe was approved by the MSSM IRB (#07-0529) and all BioMe participants provided written informed consent. Patient information was de-identified in accordance with HIPAA’s “safe harbor” method prior to access. Individuals were genotyped on the Illumina Global Screening Array; quality control (QC), imputation of the genotype data, and genetically inferred ancestry assignment for Bio*Me* is described elsewhere^23,24^.

**MGB Biobank.** ICD-9/10 codes were obtained for 29,885 individuals participating in the MGB (formerly Partners Healthcare) Biobank^25^. Patients included met a minimum data floor criteria of at least 3 visits after 2005, each more than 30 days apart, with at least one clinical note of any kind. DNA was isolated from blood and standard quality control procedures, including ancestry assignment, were performed on the genomic data. The MGB IRB (protocol #2018P002642) approved this study, and all participants provided written informed consent.
